# A systematic review on the incidence of influenza viruses in wastewater matrices: Implications for Public Health

**DOI:** 10.1101/2023.09.12.23295340

**Authors:** Mbasa Dlamini, Luyanda Msolo, Kingsley Ehi Ebomah, Nolonwabo Nontongana, Anthony Ifeanyi Okoh

## Abstract

Influenza has historically been and remains a significant global health concern, causing substantial illness worldwide. Influenza is a serious public health issue in both industrialized and developing nations and it is characterized as an acute respiratory illness resulting from infection with influenza virus. Influenza viruses are RNA viruses with a negative sense and enveloped structure. These viruses possess segmented genomes, with influenza A and B viruses being the prevalent types affecting human populations. These viruses have been associated with disease outbreaks in some regions of the world as a result of excrement being introduced into the environment. Given the global implications of influenza and the limited availability of data for many countries, particularly in the African region where the prevalence and incidence of influenza remain largely unknown, there is a lack of published information regarding the detection of influenza viruses. Therefore, the purpose of this paper is to examine or review the frequency of influenza virus detection in wastewater samples, serving as an initial step toward gaining a better understanding of the burden of influenza. This includes exploring its epidemiology, the consequences of severe influenza infections, and the development of strategies to enhance supportive care and virus-specific therapies in resource-constrained, low-income settings.

## 1. Introduction

Influenza viruses are recognized as the primary causative agents of respiratory tract diseases, resulting in significant health and socioeconomic effects globally (Marco *et al.,* 2020). These viruses are negative sense, enveloped RNA viruses that contain segmented genomes and are categorized into four types, namely influenza A, B, C, and D. All four types of influenza viruses are considered endemic, with types A and B being the most widespread and responsible for causing influenza, commonly known as the flu. Symptoms of the flu typically include chills, fever, headache, sore throat, and muscle pain (Brussow, 2021). In severe instances, influenza can lead to pneumonia, which can be particularly life-threatening for vulnerable populations such as children and the elderly.

In humans, influenza A and B viruses are the most prevalent and impose a significant burden on the global population (Caini *et al.,* 2019). Transmission occurs through respiratory droplets released during coughing, talking, and sneezing, as well as through contact with contaminated surfaces followed by touching the nose or eyes (Lin *et al.,* 2022). In terms of public health impact, influenza A and B viruses pose the greatest threat, causing seasonal epidemics that affect millions of individuals (Nickol and Kindrachuk, 2019). While most influenza virus infections are limited to the upper respiratory tract, some cases can be severe and give rise to complications such as pneumonia, acute respiratory distress syndrome, and central nervous system involvement (Ma *et al.,* 2020). Numerous studies have drawn attention to the association of influenza A with conditions like encephalitis, febrile seizures, and Guillain-Barre syndrome (Marco *et al.,* 2020; Beghi *et al.,* 2020; Gusev *et al.,* 2021; Meidaninikjeh *et al.,* 2022).

Influenza viruses differ from double-stranded DNA viruses as they consist of single-stranded RNA. Over time, these viruses undergo genetic changes and continuously evolve. This evolution occurs in two processes such as antigenic drift and antigenic shift (Hannoun, 2013). Antigenic drift is the progressive accumulation of genetic mutations within viral genes that are responsible for generating surface proteins recognized by host antibodies (Naeem *et al.,* 2020). These changes, also referred to as drift, result in minor alterations in the virus’s surface proteins, namely hemagglutinin and neuraminidase, which are antigens capable of stimulating an immune response, including antibody development, against the virus (Nooraei *et al.,* 2002). Antigenic drift occurs as influenza viruses replicate within a host, leading to ongoing modifications over time. However, antigenic shift is of greater concern compared to antigenic drift (Yewdell, 2021). Antigenic shift can give rise to an influenza virus variant that evades existing antibodies in the population. This can result in the occurrence of an influenza pandemic (Krammer, 2019). Antigenic shift refers to a significant genetic alteration in an infectious agent, resulting in a substantial modification in the antigenic protein. This modification triggers the immune systems of humans and other animals to produce antibodies. For example, an antigenic shift occurs when there is a sudden and drastic change in hemagglutinins of influenza type A viruses.

The influenza virus undergoes various stages throughout its life cycle such as entry into the host cell, entry of viral ribonucleoproteins (vRNPs) into the nucleus, viral genome transcription and replication, export of vRNPs from the nucleus, and assembly and budding at the host cell plasma membrane. This process culminates in the formation of infectious virus particles which are subsequently released into the extracellular environment, allowing them to infect new cells (Gill *et al.,* 2019). These stages are depicted by the sourced diagrams (Figures 2 and 3).

**Figure 2:**
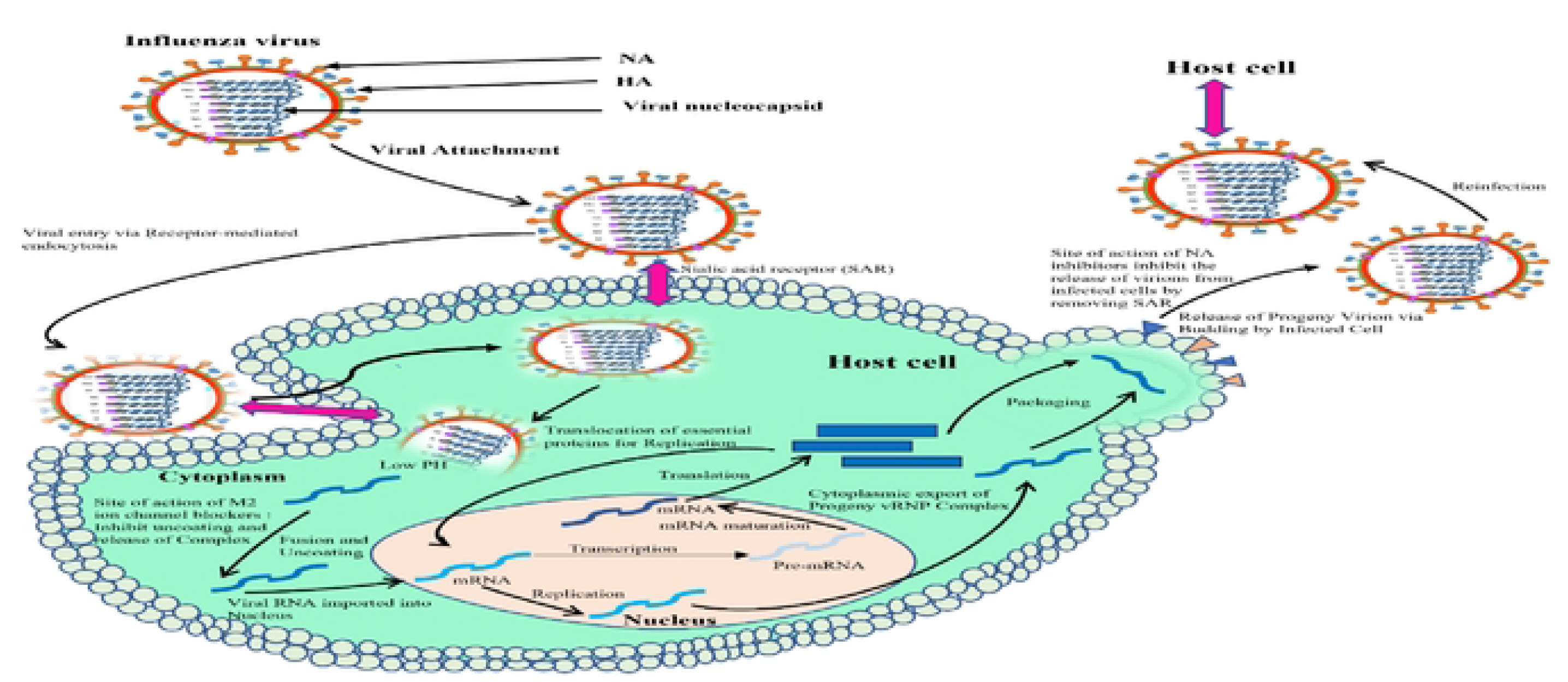
A Pictorial Representation of Influenza Virus Lifecycle.

**Figure 3:**
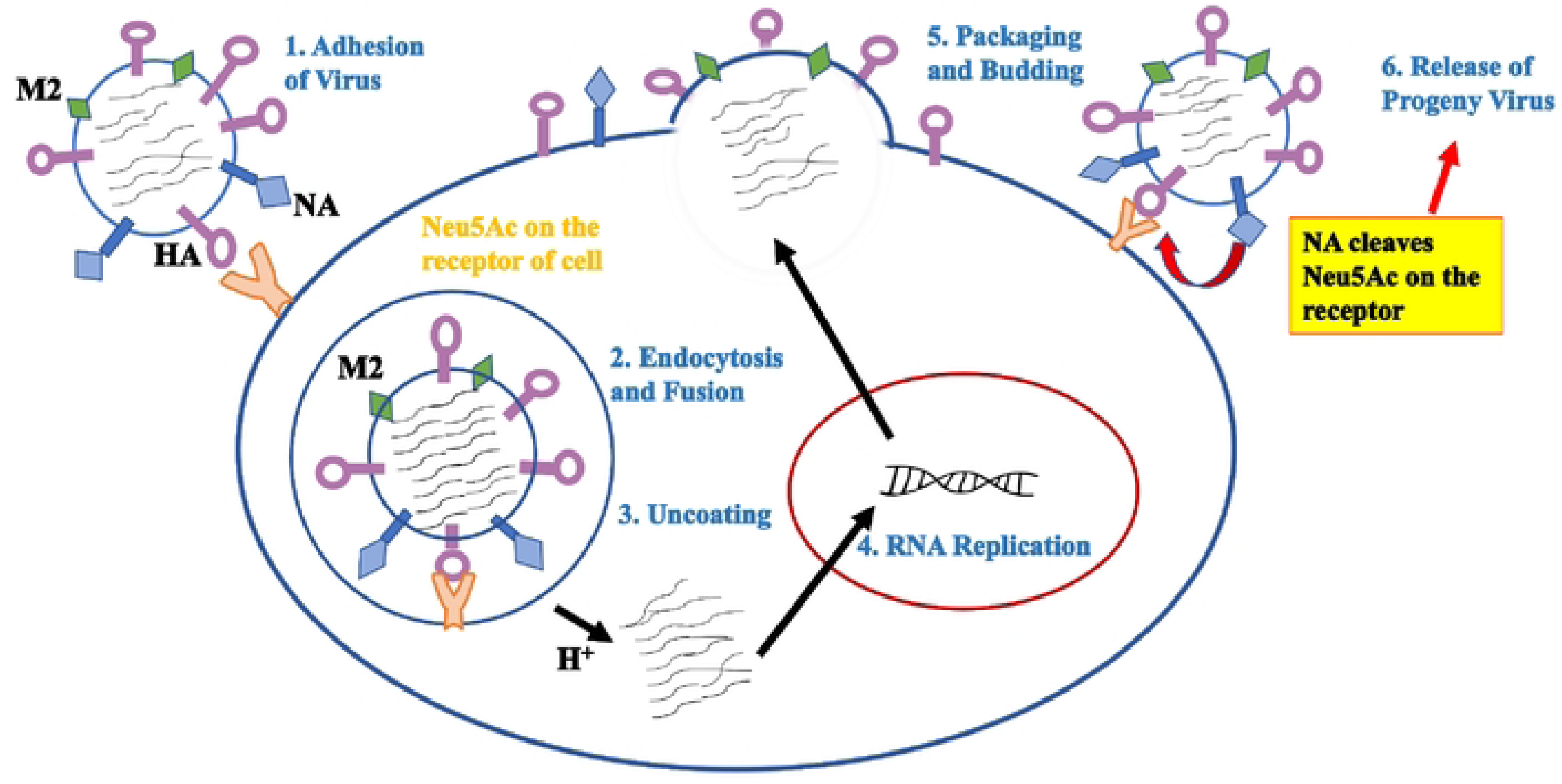
Illustrative depiction showing the lifecycle of the influenza virus.

Wastewater management plays a crucial role in controlling the spread of influenza viruses and other pathogens. There are some key issues related to wastewater management and the circulation of influenza viruses around the world. Firstly, inadequate sanitation infrastructure, for example, many regions, especially in developing countries, lack proper sanitation infrastructure, including sewage treatment plants and wastewater treatment facilities (Wang *et al.,* 2018). This can lead to the direct discharge of untreated or partially treated wastewater into rivers, lakes, or coastal areas, potentially contaminating water sources and increasing the risk of spreading influenza viruses. Urbanization and Overcrowding-rapid urbanization and population growth have put significant strain on existing wastewater infrastructure in many cities (Habib, 2022). Overcrowding can lead to inadequate sanitation facilities, leading to the improper management of wastewater. This can contribute to the spread of influenza viruses among densely populated communities. Thirdly agricultural runoff, for example in rural areas, agricultural activities can contribute to the contamination of water bodies with influenza viruses. The use of untreated or poorly treated wastewater for irrigation or the improper disposal of manure can introduce pathogens into the environment, which can contaminate water sources and potentially spread influenza viruses (Ali *et al.,* 2021).

Therefore, wastewater is considered a reservoir of a variety of human pathogens such as influenza viruses that can impact human health and contribute to a range of environmental and health complications (Shaheen, 2022). These viruses can enter the water cycle when they are expelled from the fecal matter of infected individuals. (Gerba and Pepper, 2019). The two primary goals of treating wastewater are to prevent the contamination of water sources and to maintain public health by preventing the spread of illnesses caused by influenza viruses through the protection of water resources. Henceforth, this paper is aimed at reviewing the effects on both the environment and human health caused by the prevalence of influenza viruses in untreated or inadequately treated wastewater or wastewater matrices. Addressing these issues requires a comprehensive approach that includes improving sanitation infrastructure, upgrading wastewater treatment facilities, promoting proper hygiene practices, and raising awareness about the importance of responsible wastewater management. By implementing effective wastewater management practices, the risk of spreading influenza viruses and other pathogens can be significantly reduced, protecting public health and the environment.

### 2.1 Epidemiological evidence of human health risks associated with influenza viruses

The flu is a highly contagious viral illness that can infect people across all age groups. However, certain groups are more vulnerable to its impact, and during pandemics, epidemics, and sporadic outbreaks, it has been associated with significant mortality rates (Miron *et al.,* 2021). Pregnant women, children below the age of five (especially those under the age of two), the elderly, and those undergoing chemotherapy and taking steroids are at a heightened risk of developing severe illness or complications if infected (Javanian *et al.,* 2021).

Individuals can become infected with and transmit the Influenza A virus through various means, including direct contact with bodily secretions, inhalation of respiratory droplets, and indirect contact with contaminated objects. This virus is notorious for its significant rates of illness and death, and it is responsible for both seasonal outbreaks and notable historical influenza pandemics (Bailey *et al.,* 2018). On the other side, human infections with influenza B viruses pose a continuing concern to public health. According to Javanian *et al*. (2021), direct contact with an infected individual is the main risk factor for human influenza B infection. For instance, large quantities of viral particles that are harmful to humans are expelled from infected individuals, which are then introduced into wastewater and later cause waterborne infections all over the world (Ahmad *et al.,* 2021).

Growing concerns have arisen in light of notable viral outbreaks in recent years, underscoring the potential for a severe viral pandemic originating from influenza viruses present in wastewater matrices. As highlighted by Huremovic (2019), the public’s apprehension is well-founded, considering the catastrophic impact that viral pandemics can have. Notably, historical records indicate that influenza pandemics killed people in absolute numbers than any other disease outbreak in recorded history. Another example, looking at the results of the detection of influenza A and B in the influent of two WWTPs in Germany represents how wastewater matrices are regarded as a reservoir of a variety of influenza viruses and a major cause of global concern (Dumke *et al*., 2022). Based on the findings of the study, it was proven that two German cities’ municipal wastewater contained respiratory virus RNA. For example, influenza B virus was detected in 36.0% and 57.7% of the sampled wastewater. It was concluded that more research should be done using more samples from various places to learn more or expand knowledge about the prevalence of influenza in wastewater matrices. Particularly in situations where preventive measures to mitigate the transmission of respiratory viral infections are lacking, it becomes crucial to examine and comprehend the broader implications of influenza viruses present in wastewater.

During the spring of 2022, The City of Ottawa and its neighborhoods experienced an unusual spike in influenza virus activity, which allowed for the collection of wastewater samples containing influenza viruses (Mercier *et al.,* 2022). The findings of the study were then applied to improve a procedure for measuring influenza virus RNA in wastewater. Their improved method was then used to measure influenza virus RNA in wastewater from the whole city of Ottawa, Ontario, Canada, as well as from three different neighborhoods. Influenza A virus was demonstrated in 10.3% of the samples from treatment plant 1 (WRRF), 3.1% from treatment plant 2, and 0.1% of the samples from plant 3. These results concur with those of another research that identified the influenza A virus in wastewater matrices (Wolfe *et al.,* 2022). Therefore, based on the findings, the processing of wastewater samples and detection of influenza viral signals was performed and it was concluded that wastewater indeed represents a reservoir of numerous pathogens.

To examine the potential of influenza transmission channels, several investigations of the influenza virus in wastewater have been conducted. Few research has described a procedure for successfully detecting influenza virus in wastewaters up to this point (Wurtzer *et al.,* 2021). For example, a recent study showed a good correlation between wastewater measurements of influenza A viruses and reported clinical IAV cases seen as part of monitoring systems for athletes at Stanford University and Michigan University’s campus (Mercier *et al.,* 2022). However, there is a dearth of knowledge on the measurement, trends, and, most critically, the relationship between the influenza virus signal in wastewater in cities and neighborhood communities. Therefore, new information linked to influenza WWS is thus necessary to clarify other characteristics of the influenza virus in wastewater matrices. According to (Schwab and Malleret, 2020), wastewater surveillance for influenza possesses several advantageous characteristics, including its ability to provide anonymous, aggregated, cost-effective, and rapid monitoring of a substantial portion of communities. Due to its involuntary contributions, WWS serves as an important tool for public health units or agencies.

**Figure 4:**
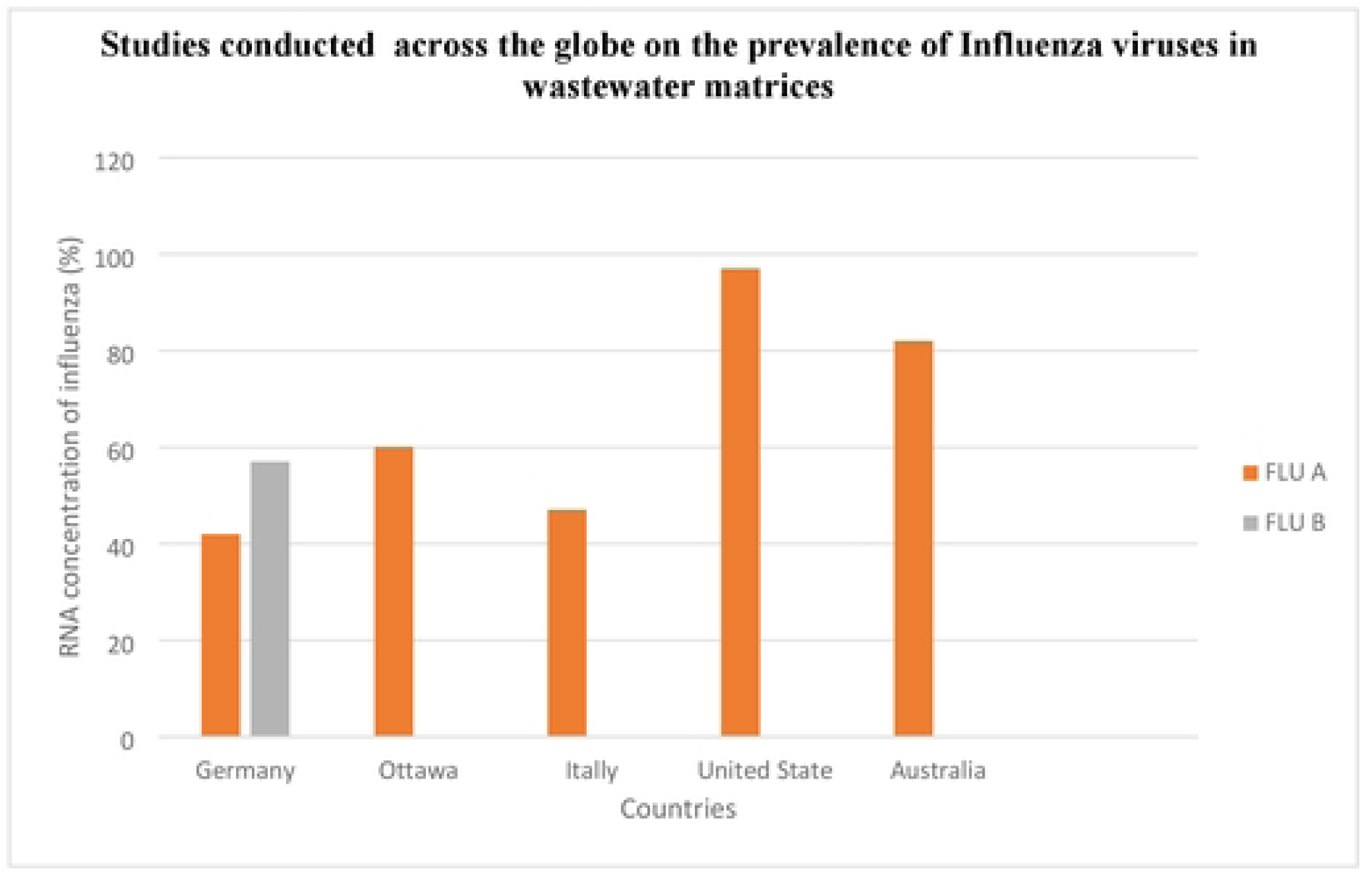
Studies conducted across the globe on the prevalence of influenza viruses associated with wastewater.

**Table 1:** Most productive authors in research related to the prevalence of Influenza viruses in wastewater milieu from 2012 to 2022 (Scopus and google scholar)

### 2.2 How wastewater treatment helps prevent disease

Wastewater treatment is the process of removing, eliminating, or inactivating the majority of contaminants and disease-causing organisms, such as influenza viruses from wastewater (Anand *et al.,* 2020). Globally, wastewater poses significant risks of infections, which accounts for around a million fatalities each year, nearly half of which are children and more than 90% of which occur in underdeveloped nations. The main cause of these fatalities is ingesting fecal pathogens from people or animals (Javanian *et al.,* 2021). Hence, there is a desire among researchers to create and examine criteria based on a clearer framework for determining the danger of influenza viruses in wastewater. A technique known as wastewater-based epidemiology (WBE), which is a tool used to estimate the likelihood of infectious diseases among individuals exposed to materials associated with waste, is becoming more popular as a result of this aim. According to Lin *et al.,* (2022), this tool, enables the identification and surveillance of both pandemic and seasonal influenza outbreaks.

#### 2.2.1 Treatment and control measures

The WHO and the National Institutes of Health have acknowledged the enormous worldwide burden of infectious disease epidemics caused by the influenza virus and the necessity to build prediction and preventive systems (Xagoraraki *et al.,* 2020). According to Korkmaz *et al.,* (2021), suggest that immunization is the most effective measure to prevent influenza infection and its sequelae.

Antiviral drugs play a crucial role in supporting the body’s natural defenses against specific viruses that cause diseases. These medications are recognized as essential tools in the prevention and containment of viruses (Peterson *et al.,* 2020). Individuals who are at high risk for flu-related complications, such as older adults or those with weakened immune systems, should seek antiviral treatment as early as possible. Early administration of antiviral medication can result in a milder illness, preventing the need for hospitalization in vulnerable individuals susceptible to severe flu complications (Kausar *et al.,* 2021). Moreover, certain studies suggest that early use of antiviral drugs can reduce the risk of mortality among hospitalized individuals with influenza.

Currently, there are three recommended antiviral medications, namely oseltamivir, zanamivir, and peramivir, for the treatment of influenza. These drugs act by bolstering the immune system’s ability to combat viral infections and reducing the viral load present in the body (Anand *et al.,* 2020). Rapid diagnosis and treatment are crucial, as antiviral drugs can effectively treat both influenza A and B infections when administered within 48 hours of the onset of flu symptoms. For instance, oseltamivir, an antiviral drug commonly used, specifically targets and treats both influenza A and B viruses. It alleviates flu-related symptoms, resulting in a reduction in severity and a shortened recovery period of one to two days (Kalra *et al.,* 2011). These symptoms include stuffy nose, coughing, sore throat, fever or chills, pains, and fatigue. Oseltamivir functions by targeting the influenza virus, impeding its proliferation within the body, and alleviating flu symptoms.

### 2.3 Environmental factors influencing the transmission of the influenza viruses

Several factors influence the severity and transmission of influenza, including the natural and acquired hosts, viral-host interactions, environmental persistence, virus stability, and anthropogenic interventions (Bai *et al.,* 2021). Variations in temperature, humidity, pH, salinity, air pollution, and solar radiation can have an impact on a virus’ ability to survive in varied habitats. In temperate regions with higher latitudes, influenza tends to thrive and spread more effectively in cool and dry conditions, whereas, in tropical and subtropical areas characterized by humid and rainy climates, outbreaks are more prevalent, as observed by Sooryanarain and Elankumaran (2015).

Viruses that target the human respiratory system and give rise to diseases possess diverse mechanisms of transmission. However, they all share the capability to spread among individuals. The transmissibility of these viruses is influenced by the circumstances in which a pathogen and a host come into contact. Various hypotheses have been proposed to elucidate the intricate relationship between temperature, humidity, and the notable seasonality of viruses (Moriyana *et al.,* 2020). These theories encompass alterations in host behavior and adjustments in the virus’s susceptibility to infection and stability under different environmental conditions. High temperatures can cause the denaturation of viral capsid proteins’ secondary structures, while cold temperatures can lead to nucleic acid degradation, for example, disease prevalence tends to decrease as the temperature rises Wolkoff *et al.,* (2021)

The transmissibility of viruses can be impacted by ambient humidity, which has implications not only for the stability of the virus but also for the size of respiratory droplets as the water content evaporates. The size of these droplets, in turn, plays a role in determining whether they settle quickly to the ground or remain airborne long enough to be inhaled by a susceptible host (Wang *et al.,* 2021). Moreover, the pH levels can influence the transmission of influenza viruses. For instance, when the pH reaches a certain acidic threshold, it triggers a conformational change in the HA protein, exposing the fusion peptide. This critical process initiates the fusion of the virus with the host, leading to the uncoating and the release of the viral genome into the host cell’s cytoplasm (Gill *et al.,* 2019).

**Figure 5:**
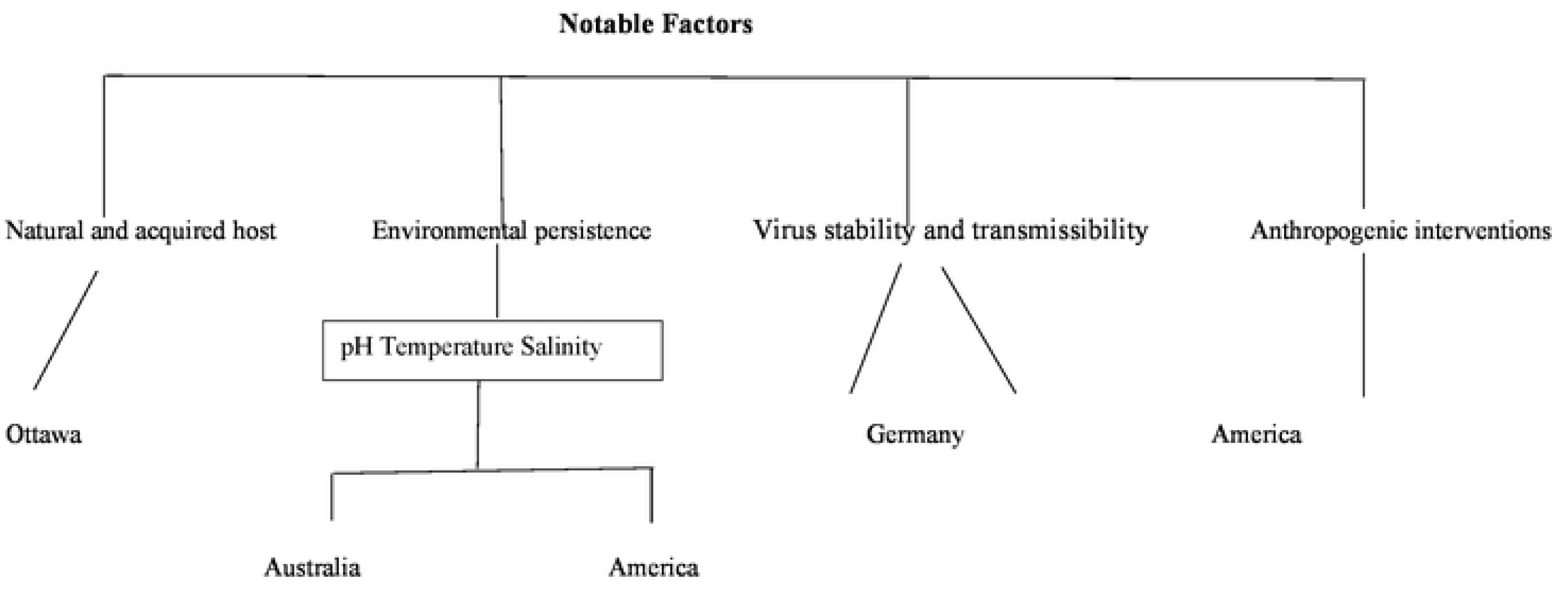
A dendrogram that illustrates some of the notable factors that influence the transmission of influenza viruses in different regions.

### 2.4 Pathogenesis of influenza viruses

Influenza viruses are known to regularly circulate within human population worldwide, giving rise to seasonal epidemics (Petrova and Russell, 2018). As they circulate, these viruses undergo gradual mutations through a process referred to as antigenic drift. Upon respiratory transmission, the virus attaches itself to respiratory epithelial cells in the trachea and bronchi, subsequently penetrating them. This leads to viral replication, destroying the host cell (Wang *et al.,* 2021).

The pathogenicity of the influenza virus relies on the interplay between viral proteins and host immune responses, encompassing both innate and acquired immunity. This highlights the significance of both viral factors and the host immune system in determining the course of influenza pathogenesis. The immune system serves as a defense mechanism against influenza virus infection. When respiratory epithelial cells or alveolar macrophages are infected by IAV, the single-stranded RNA of the virus is recognized by toll-like receptor (TLR) 7 and retinoic acid-inducible gene-I (Wu *et al.,* 2015). Activation of the TLR7 and RIG-I signaling pathways leads to the production of type I interferons and triggers antiviral responses in the host (Moriyama and Ichinohe, 2019).

## 3. Material and methods

The study followed the updated Preferred Reporting Items for Systematic Reviews and Metal Analyses extension for PRISMA guidelines (Page *et al.,* 2021).

### 3.1 Study objective

The objective of this study was to investigate the frequency of detection of influenza viruses in wastewater matrices.

### 3.2 Systematic review

The following inclusion and exclusion criteria were used when selecting the studies to be included in a systematic review.

#### 3.2.1 Inclusion criteria

Studies that measure Influenza viruses in wastewater were included in this study. Articles were limited to those published in English and those conducted in communities or populations, including private/public facilities such as hospitals connected to sewer lines. The studies which provide intelligible information on the method of detecting influenza viruses using: real-time reverse transcriptase quantitative polymerase chain reaction (RT-qPCR) was considered for inclusion in the systematic review. Studies that reported data based on the following metrics were also included: (1) detection of Influenza viruses from wastewater treatment plants (WWTPs); (2) linking data obtained from the WWTPs to RNA prevalence, resulting in an estimated value for community infection.

#### 3.2.2 Exclusion criteria

Summarized publications, abstracts, subscription-based articles without access, newspaper reports and studies using Influenza viruses in other matrices or contexts other than in wastewater, were excluded from this review.

#### 3.2.3 Search strategy

Searches were performed using Google Scholar and Scopus databases using the keywords mentioned and a combination of the keywords. Other website such as the World Health Organisation library databases was also used to search for data. The keywords used were based on the terminology normally used in this subject, including: “Wastewater” OR “Epidemiology” AND “Influenza” OR “Viruses”. The searched articles were screened from their titles, abstracts, and full text to select the eligible studies.

### 3.3 Methods of review

#### 3.3.1 Data extraction

Data was extracted from google scholar and scopus databases by reviewing titles, abstracts for inclusion and exclusion criteria.

#### 3.3.2 Data synthesis

A systematic review and meta-analyses were done. Therefore, as meta-analyses was performed statistical heterogeneity and publication bias were not assessed.

### 3.4 Quality assessment

Included articles were assessed for methodologies bias using the previously described tool by Wolfe *et al.,* 2022. Items on the tool include the following characteristics: Clarity of sampling approach; consistent use of proper processing for virus concentration; method of virus extraction and detection; presence of viral quantification process, clarity on sampling site(s) and transparency in results (including statistics) presentation.

## 4. Results

### 4.1 Description of studies

Our keyword search found 418 potential articles (68 from Scopus and 350 from google scholar). Following further screening, 68 reports were excluded. 20 articles (5 from scopus and 15 from google scholar) were used for data extraction.

### 4.2 Study characteristics

Twenty articles (5 from scopus and 15 from google scholar) were reviewed for qualitative synthesis. Summarized characteristics from each article: authors, country, population size, number of samples collected and PCR methods and targeted genes were used (Table 2). Most studies were conducted in developed countries, whereas only one study was conducted in the African region. Municipal wastewater treatment facilities were primarily used to perform the studies, whereas some studies chose a defined community, such as hospitals, universities, or neighbourhoods.

## 5. Discussion

Twenty studies (5 from scopus and 15 from google scholar) were selected in the current review. Our review provides some insights into the potential risks posed by influenza viruses detected in wastewater matrices which serve as an initial step toward gaining a better understanding of the burden of influenza in the environment. This includes exploring its epidemiology, the consequences of severe influenza infections and by preventing the spread of illnesses caused by influenza viruses through the fortification of water resources

Monitoring influenza viruses in wastewater matrices is a crucial epidemiological and public health model with a number of essential advantages such as (1) Early detection and Monitoring-Wastewater surveillance enables the early detection and frequency of influenza viruses in a community, prior to the reporting of clinical cases. This can assist public health authorities track changes over time and can provide valuable insights into the prevalence of the virus in a population (Sim *et al.,* 2020). (2) Wastewater monitoring offers a population-level perspective, enabling a more thorough evaluation of the virus’s presence throughout an area or community. This information can help public health authorities to prepare and allocate resources and timeously respond effectively to outbreaks. (3) Early Warning System: Monitoring wastewater can act as an early warning system for epidemics that can occur, for example increased public health testing and treatments may be required in the affected region if a significant increase in viral RNA is detected in wastewater. (4) Tracking Variants: Influenza viruses have the ability to mutate and may have distinct characteristics, for example influenza can undergo small genetic changes, called point mutations, which can result in slightly different strains (Shepard *et al.,* 2016).

Therefore, wastewater surveillance can help in tracking the prevalence of different influenza strains and variants in a community, aiding in vaccine development and distribution strategies (Lin *et al.,* 2022). In summary, surveilling influenza viruses in wastewater matrices is a valuable tool for public health authorities and researchers. It offers population-level data, cost-effectiveness, and the ability to track variants, ultimately aiding in the prevention and control of influenza outbreaks.

The scarcity of studies conducted in the African region on the prevalence of influenza viruses in wastewater matrices can be attributed to several factors such as limited infrastructure, data collection challenges, limited research collaboration, low awareness and seasonal nature of influenza (Shaheen *et al.,* 2020). For example, limited infrastructure: Many African countries face challenges in terms of infrastructure and resources for scientific research, including wastewater surveillance. Lack of financial assistance, laboratory facilities and trained personnel can hinder the ability to conduct such studies. Data collection challenges: collecting and analyzing wastewater samples for influenza surveillance requires specialized equipment and expertise. In many African countries, these capabilities may be lacking or underdeveloped. Low awareness: Influenza surveillance in wastewater is a relatively new concept, and there may be a lack of awareness or understanding of its potential benefits among public health officials and researchers in the African region. Seasonal nature of influenza: Influenza infections tends to be seasonal in many regions, including Africa, which can make it more difficult to conduct year-round surveillance and may limit the perceived urgency of such studies (Moriyama *et al.,* 2020).

Therefore, to address the scarcity of studies in the African region, it is essential to prioritize capacity building, promote international collaboration, increase awareness of the importance of wastewater surveillance, and allocate resources specifically for this purpose. Investing in research infrastructure and training for local researchers and institutions can help bridge the gap and contribute to a more comprehensive understanding of influenza viral infections in the region. Additionally, raising awareness about the potential benefits of wastewater surveillance for public health can encourage greater research efforts in this area.

## Conclusion

Influenza viruses continue to pose a significant burden on the global population, leading to ongoing challenges in the healthcare system and raising concerns about the increasing frequency of epidemics worldwide. To the best capacity of our knowledge, many of the variables and factors involved in the transmission of infectious diseases through wastewater are limited and often unpredictable. Additionally, there are significant gaps regarding the potential role of wastewater matrices in spreading respiratory viruses, specifically influenza viruses. To address these issues, it is crucial for environmental engineers, medical virologists, and public health researchers to collaborate and develop effective methods for treating influenza viruses present in complex sample matrices like wastewater. This necessitates implementing a treatment process that includes site restrictions to reliably reduce the concentration of pathogens to a safe level before human or animal contact. In conclusion, there is a pressing need for surveillance techniques or tools that can accurately estimate the number, epidemiology, and distribution of influenza viruses. Such measures are essential for timely intervention and to prevent the further spread of the disease.

## Data Availability

N/A

## Acknowledgment

We are grateful to the South African Medical Research Council and National Research Foundation for financial support.

## References

Anand, A., Rajchakit, U. and Sarojini, V., 2020. Detection and removal of biological contaminants in water: the role of nanotechnology. In Nanomaterials for the Detection and Removal of Wastewater Pollutants (pp. 69-110). Elsevier.

Bai, Y., Jones, J.C., Wong, S.S. and Zanin, M., 2021. Antivirals targeting the surface glycoproteins of influenza virus: Mechanisms of action and resistance. Viruses, 13(4), p.624.

Bailey, E.S., Choi, J.Y., Fieldhouse, J.K., Borkenhagen, L.K., Zemke, J., Zhang, D. and Gray, G.C., 2018. The continual threat of influenza virus infections at the human–animal interface: what is new from a one health perspective? Evolution, medicine, and public health, 2018(1), pp.192–198.

Beghi, E., Feigin, V., Caso, V., Santalucia, P. and Logroscino, G., 2020. COVID-19 infection and neurological complications: present findings and future predictions. Neuroepidemiology, 54(5), pp.364–369.

Bi, C., Ramos-Mandujano, G., Tian, Y., Hala, S., Xu, J., Mfarrej, S., Esteban, C.R., Delicado, E.N., Alofi, F.S., Khogeer, A. and Hashem, A.M., 2021. Simultaneous detection and mutation surveillance of SARS-CoV-2 and multiple respiratory viruses by rapid field-deployable sequencing. Med, 2(6), pp.689–700.

Boehm, A.B., Hughes, B., Duong, D., Chan-Herur, V., Buchman, A., Wolfe, M.K. and White, B.J., 2022. Wastewater surveillance of human influenza, metapneumovirus, parainfluenza, respiratory syncytial virus (RSV), rhinovirus, and seasonal coronaviruses during the COVID-19 pandemic. medRxiv, pp.2022–09.

Brisebois, E., Veillette, M., Dion-Dupont, V., Lavoie, J., Corbeil, J., Culley, A. and Duchaine, C., 2018. Human viral pathogens are pervasive in wastewater treatment center aerosols. Journal of Environmental Sciences, 67, pp.45–53.

Brüssow, H, 2021. Clinical evidence that the pandemic from 1889 to 1891 commonly called the Russian flu might have been an earlier coronavirus pandemic. Microbial Biotechnology, 14(5), pp.1860–1870.

Caini, S., Kusznierz, G., Garate, V.V., Wangchuk, S., Thapa, B., de Paula Júnior, F.J., Ferreira de Almeida, W.A., Njouom, R., Fasce, R.A., Bustos, P. and Feng, L., 2019. The epidemiological signature of influenza B virus and its B/Victoria and B/Yamagata lineages in the 21st century. PloS one, 14(9), p.e0222381.

Chauhan, R.P. and Gordon, M.L., 2022. An overview of influenza A virus genes, protein functions, and replication cycle highlighting important updates. Virus Genes, 58(4), pp.255–269.

Dumke, R., Geissler, M., Skupin, A., Helm, B., Mayer, R., Schubert, S., Oertel, R., Renner, B. and Dalpke, A.H., 2022. Simultaneous detection of SARS-CoV-2 and influenza virus in wastewater of two cities in Southeastern Germany, January to May 2022. International journal of environmental research and public health, 19(20), p.13374.

Gaidet, N., Caron, A., Cappelle, J., Cumming, G.S., Balança, G., Hammoumi, S., Cattoli, G., Abolnik, C., Servan de Almeida, R., Gil, P. and Fereidouni, S.R., 2012. Understanding the ecological drivers of avian influenza virus infection in wildfowl: a continental-scale study across Africa. Proceedings of the Royal Society B: Biological Sciences, 279(1731), pp.1131–1141.

Gerba, C.P. and Pepper, I.L., 2019. Microbial contaminants. Environmental and pollution science, pp.191–217.

Ghernaout, D., Elboughdiri, N. and Al Arni, S., 2020. New insights towards disinfecting viruses– short notes. Journal of Water Reuse and Desalination, 10(3), pp.173–186.

Gill, S., Catchpole, R. and Forterre, P., 2019. Extracellular membrane vesicles in the three domains of life and beyond. FEMS microbiology reviews, 43(3), pp.273–303.

Gusev, E.I., Martynov, M.Y., Boyko, A.N., Voznyuk, I.A., Latsh, N.Y., Sivertseva, S.A., Spirin, N.N. and Shamalov, N.A., 2021. The novel coronavirus infection (COVID-19) and nervous system involvement: mechanisms of neurological disorders, clinical manifestations, and the organization of neurological care. Neuroscience and Behavioral Physiology, 51, pp.147–154.

Huremović, D., 2019. A brief history of pandemics (pandemics throughout history). Psychiatry of pandemics: a mental health response to infection outbreak, pp.7–35.

Javanian, M., Barary, M., Ghebrehewet, S., Koppolu, V., Vasigala, V. and Ebrahimpour, S., 2021. A brief review of influenza virus infection. Journal of Medical Virology, 93(8), pp.4638–4646.

Kalra, M., Khatak, M. and Khatak, S., 2011. Cold and flu: conventional vs. botanical and nutritional therapy. Int J Drug Dev Res, 3(1), pp.314–327.

Kaplan, G.G., Tang, W., Banerjee, R., Adigopula, B., Underwood, F.E., Tanyingoh, D., Wei, S.C., Lin, W.C., Lin, H.H. and Li, J., 2019. Population density and risk of inflammatory bowel disease: a prospective population-based study in 13 countries or regions in Asia-Pacific. Official journal of the American College of Gastroenterology| ACG, 114(1), pp.107–115.

Kausar, S., Said Khan, F., Ishaq Mujeeb Ur Rehman, M., Akram, M., Riaz, M., Rasool, G., Hamid Khan, A., Saleem, I., Shamim, S. and Malik, A., 2021. A review: Mechanism of action of antiviral drugs. International journal of immunopathology and pharmacology, 35, p.20587384211002621.

Kevill, J.L., Lambert-Slosarska, K., Pellett, C., Woodhall, N., Pântea, I., Alex-Sanders, N. and Jones, D.L., 2022. Assessment of two types of passive sampler for the efficient recovery of SARS-CoV-2 and other viruses from wastewater. Science of the Total Environment, 838, p.156580.

Kikkert, M., 2020. Innate immune evasion by human respiratory RNA viruses. Journal of innate immunity, 12(1), pp.4–20.

Korkmaz, N., Nazik, S., Gümüştakım, R.Ş., Uzar, H., Kul, G., Tosun, S., Torun, A., Demirbakan, H., Seremet Keskin, A., Kaçmaz, A.B. and Erdem, H.A., 2021. Influenza vaccination rates, knowledge, attitudes and behaviours of healthcare workers in Turkey: A multicentre study. International Journal of Clinical Practice, 75(1), p.e13659.

Krammer, F., 2019. The human antibody response to influenza A virus infection and vaccination. Nature Reviews Immunology, 19(6), pp.383–397.

Lin, N., Servetas, S., Jackson, S., Lippa, K., Parratt, K., Mattson, P., Beahn, C., Mattioli, M., Gutierrez, S., Focazio, M. and Smith, T., 2022. Report on the DHS/NIST Workshop on Standards for an Enduring Capability in Wastewater Surveillance for Public Health (SWWS Workshop). In NIST Special Publication. National Institute of Standards and Technology.

Ma, X., Liang, M., Ding, M., Liu, W., Zhou, X. and Ren, H., 2020. Extracorporeal membrane oxygenation (ECMO) in critically ill patients with coronavirus disease 2019 (COVID-19) pneumonia and acute respiratory distress syndrome (ARDS). Medical science monitor: international medical journal of Experimental and clinical research, 26, pp.e925364–1.

Marco, C., Massimo, C., Alessandro, T., Wen-Can, J., Cheng-Bin, W. and Sergio, B., 2020. The COVID-19 pandemic. Critical Reviews in Clinical Laboratory Sciences, 57(6), pp.365–388

Meidaninikjeh, S., Sabouni, N., Taheri, M., Borjkhani, M., Bengar, S., Majidi Zolbanin, N., Khalili, A. and Jafari, R., 2022. SARS-CoV-2 and Guillain–Barré Syndrome: lessons from viral infections. Viral Immunology, 35(6), pp.404–417.

Mercier, E., D’Aoust, P.M., Thakali, O., Hegazy, N., Jia, J.J., Zhang, Z., Eid, W., Plaza-Diaz, J., Kabir, M.P., Fang, W. and Cowan, A., 2022. Municipal and neighborhood-level wastewater surveillance and subtyping of an influenza virus outbreak. Scientific Reports, 12(1), p.15777.

Moriyama, M. and Ichinohe, T., 2019. High ambient temperature dampens adaptive immune responses to influenza A virus infection. Proceedings of the National Academy of Sciences, 116(8), pp.3118–3125.

Moriyama, M., Hugentobler, W.J. and Iwasaki, A., 2020. Seasonality of respiratory viral infections. Annual review of virology, 7, pp.83–101.

Naeem, A., Elbakkouri, K., Alfaiz, A., Hamed, M.E., Alsaran, H., AlOtaiby, S., Enani, M. and Alosaimi, B., 2020. Antigenic drift of hemagglutinin and neuraminidase in seasonal H1N1 influenza viruses from Saudi Arabia in 2014 to 2015. Journal of Medical Virology, 92(12), pp.3016–3027.

Nickol, M.E. and Kindrachuk, J., 2019. A year of terror and a century of reflection: perspectives on the great influenza pandemic of 1918–1919. BMC infectious diseases, 19(1), pp.1–10

Nooraei, S., Bahrulolum, H., Hoseini, Z.S., Katalani, C., Hajizade, A., Easton, A.J. and Ahmadian, G., 2021. Virus-like particles: preparation, immunogenicity and their roles as nanovaccines and drug nanocarriers. Journal of nanobiotechnology, 19(1), pp.1–27.

O’Brien, E. and Xagoraraki, I., 2019. A water-focused one-health approach for early detection and prevention of viral outbreaks. One Health, 7, p.100094.

Page, M.J., McKenzie, J.E., Bossuyt, P.M., Boutron, I., Hoffmann, T.C., Mulrow, C.D., Shamseer, L., Tetzlaff, J.M., Akl, E.A., Brennan, S.E. and Chou, R., 2021. The PRISMA 2020 statement: an updated guideline for reporting systematic reviews. International journal of surgery, 88, p.105906.

Petersen, E., Koopmans, M., Go, U., Hamer, D.H., Petrosillo, N., Castelli, F., Storgaard, M., Al Khalili, S. and Simonsen, L., 2020. Comparing SARS-CoV-2 with SARS-CoV and influenza pandemics. The Lancet infectious diseases, 20(9), pp.e238–e244.

Ramos-Mandujano, G., Salunke, R., Mfarrej, S., Rachmadi, A.T., Hala, S., Xu, J., Alofi, F.S., Khogeer, A., Hashem, A.M., Almontashiri, N.A. and Alsomali, A., 2021. A robust, safe, and scalable magnetic nanoparticle workflow for RNA extraction of pathogens from clinical and wastewater samples. Global Challenges, 5(4), p.2000068.

Rönnqvist, M., Ziegler, T., von Bonsdorff, C.H. and Maunula, L., 2012. Detection method for avian influenza viruses in water. Food and environmental virology, 4, pp.26–33.

Rusiñol, M., Martínez-Puchol, S., Forés, E., Itarte, M., Girones, R. and Bofill-Mas, S., 2020. Concentration methods for the quantification of coronavirus and other potentially pandemic enveloped virus from wastewater. Current opinion in environmental science & health, 17, pp.21–28.

Schwab, K. and Malleret, T., 2020, March. The great reset. In World economic forum, Geneva (Vol. 22).

Shaheen, M.N., 2022. The concept of one health applied to the problem of zoonotic diseases. Reviews in Medical Virology, 32(4), p.e2326.

Shepard, S.S., Meno, S., Bahl, J., Wilson, M.M., Barnes, J. and Neuhaus, E., 2016. Viral deep sequencing needs an adaptive approach: IRMA, the iterative refinement meta-assembler. BMC genomics, 17, pp.1–18.

Shie, J.J. and Fang, J.M., 2019. Development of effective anti-influenza drugs: congeners and conjugates–a review. Journal of Biomedical Science, 26(1), pp.1–20.

Sieben, C., Kappel, C., Zhu, R., Wozniak, A., Rankl, C., Hinterdorfer, P., Grubmüller, H. and Herrmann, A., 2012. Influenza virus binds its host cell using multiple dynamic interactions. Proceedings of the National Academy of Sciences, 109(34), pp.13626–13631.

Silverman, A.I. and Boehm, A.B., 2021. Systematic review and meta-analysis of the persistence of enveloped viruses in environmental waters and wastewater in the absence of disinfectants. Environmental Science & Technology, 55(21), pp.14480–14493.

Sims, N. and Kasprzyk-Hordern, B., 2020. Future perspectives of wastewater-based epidemiology: monitoring infectious disease spread and resistance to the community level. Environment international, 139, p.105689

Skelton, R.M. and Huber, V.C., 2022. Comparing influenza virus biology for understanding influenza d virus. Viruses, 14(5), p.1036.

Sooryanarain, H. and Elankumaran, S., 2015. Environmental role in influenza virus outbreaks. Annu. Rev. Anim. Biosci., 3(1), pp.347–373.

Teirlinck, A.C., Van Asten, L., Brandsema, P.S., Dijkstra, F., Donker, G.A., van Gageldonk-Lafeber, A.B., Hooiveld, M., de Lange, M.M.A., Marbus, S.D., Meijer, A. and van der Hoek, W., 2017. Annual report Surveillance of influenza and other respiratory infections in the Netherlands: Winter 2016/2017.

Wang, S., Qiu, Z., Hou, Y., Deng, X., Xu, W., Zheng, T., Wu, P., Xie, S., Bian, W., Zhang, C. and Sun, Z., 2021. AXL is a candidate receptor for SARS-CoV-2 that promotes infection of pulmonary and bronchial epithelial cells. Cell research, 31(2), pp.126–140.

Wang, X., Cao, R., Zhang, H., Liu, J., Xu, M., Hu, H., Li, Y., Zhao, L., Li, W., Sun, X. and Yang, X., 2020. The anti-influenza virus drug, arbidol is an efficient inhibitor of SARS-CoV-2 in vitro. Cell discovery, 6(1), p.28.

Webster, R.G. and Govorkova, E.A., 2014. Continuing challenges in influenza. Annals of the New York Academy of Sciences, 1323(1), pp.115–139.

Wigginton, K.R., Ye, Y. and Ellenberg, R.M., 2015. Emerging investigators series: the source and fate of pandemic viruses in the urban water cycle. Environmental Science: Water Research & Technology, 1(6), pp.735–746.

Wolfe, M.K., Duong, D., Bakker, K.M., Ammerman, M., Mortenson, L., Hughes, B., Arts, P., Lauring, A.S., Fitzsimmons, W.J., Bendall, E. and Hwang, C.E., 2022. Wastewater-based detection of two influenza outbreaks. Environmental Science & Technology Letters, 9(8), pp.687–692.

Wolkoff, P., Azuma, K. and Carrer, P., 2021. Health, work performance, and risk of infection in office-like environments: The role of indoor temperature, air humidity, and ventilation. International Journal of Hygiene and Environmental Health, 233, p.113709.

Wu, W., Zhang, W., Duggan, E.S., Booth, J.L., Zou, M.H. and Metcalf, J.P., 2015. RIG-I and TLR3 are both required for maximum interferon induction by influenza virus in human lung alveolar epithelial cells. Virology, 482, pp.181–188.

Wurtzer, S., Waldman, P., Ferrier-Rembert, A., Frenois-Veyrat, G., Mouchel, J.M., Boni, M., Maday, Y., Marechal, V. and Moulin, L., 2021. Several forms of SARS-CoV-2 RNA can be detected in wastewaters: implication for wastewater-based epidemiology and risk assessment. Water Research, 198, p.117183.

Xagoraraki, Irene, and Evan O’Brien. "Wastewater-based epidemiology for early detection of viral outbreaks." Women in water quality: Investigations by prominent female engineers (2020): 75–97.

Ye, Y., Ellenberg, R.M., Graham, K.E. and Wigginton, K.R., 2016. Survivability, partitioning, and recovery of enveloped viruses in untreated municipal wastewater. Environmental science & technology, 50(10), pp.5077–5085.

Yeganeh, B., Moghadam, A.R., Tran, A.T., Rahim, M.N., Ande, S.R., Hashemi, M., Coombs, K.M. and Ghavami, S., 2013. Asthma and influenza virus infection: focusing on cell death and stress pathways in influenza virus replication. Iranian Journal of Allergy, Asthma and Immunology, pp.1–17.

Yewdell, J.W., 2021. Antigenic drift: understanding COVID-19. Immunity, 54(12), pp.2681–26

Zhang, H., Wang, Z., Chang, C., Javed, A., Ali, K., Du, W., Niazi, N.K., Mao, K. and Yang, Z., 2021. Occurrence of various viruses and recent evidence of SARS-CoV-2 in wastewater systems. Journal of hazardous materials, 414, p.125439.

