## Supplemental Figure 1 for "A systematic review on the incidence of influenza viruses in wastewater matrices: Implications for Public Health"

**Identification of studies via databases and registers**

Records removed *before screening*:

Duplicate records removed (n = 15)

Records marked as ineligible by automation tools (n = 25)

Records removed for other reasons (n = 0)

Records identified from*: Scopus (68) and Google scholar (350)

Databases (n = 418)

**Identification**

Records screened

(n = 418)

Records excluded**

(n =225 )

Reports sought for retrieval

(n = 193)

Reports not retrieved

(n = 90 )

**Screening**

Reports assessed for eligibility

(n = 100 )

Reports excluded:

Reason 1 (n =68 ) No information of interest reported on the studies

Reason 2 (n = 12 ): Reviews

Studies included in review

(n =20) 5 from scopus and 15 from google scholar

**Included**

**Figure 1**. Flow diagram of search, screening, and study selection of systematic review.

*Consider, if feasible to do so, reporting the number of records identified from each database or register searched (rather than the total number across all databases/registers).

**If automation tools were used, indicate how many records were excluded by a human and how many were excluded by automation tools.

*From:*  Page MJ, McKenzie JE, Bossuyt PM, Boutron I, Hoffmann TC, Mulrow CD, et al. The PRISMA 2020 statement: an updated guideline for reporting systematic reviews. BMJ 2021;372:n71. doi: 10.1136/bmj.n71

For more information, visit: <http://www.prisma-statement.org/>
