## Supplementary Figures 2-5 for "A systematic review on the incidence of influenza viruses in wastewater matrices: Implications for Public Health"

**Supplementary Data: Figures**

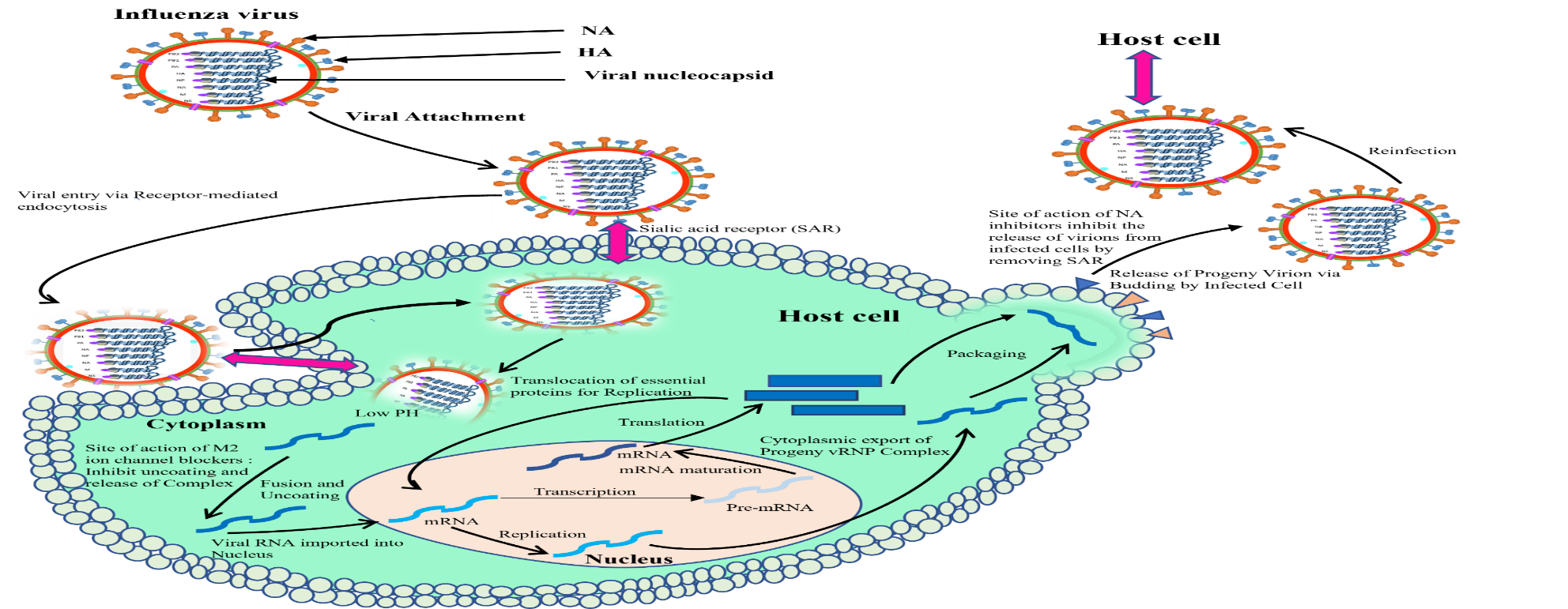

**Figure 2**: A Pictorial Representation of Influenza Virus Lifecycle

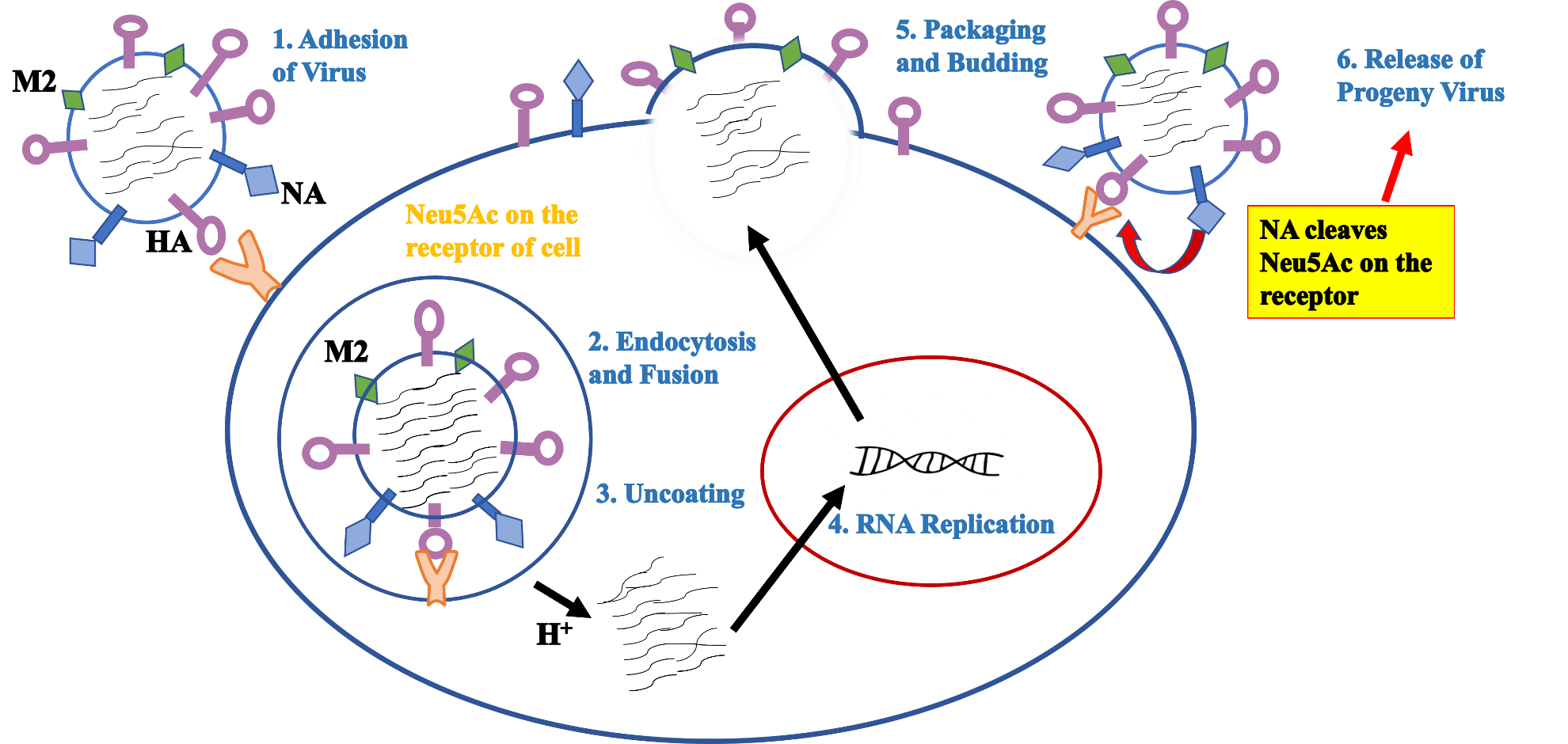

**Figure 3:** Illustrative depiction showing the lifecycle of the influenza virus

Studies conducted across the globe on the prevalence of Influenza viruses in wastewater matrices

RNA concentration of influenza (%)

| Countries | FLU A | FLU B |
| --- | --- | --- |
| Germany | 42 | 57 |
| Ottawa | 60 |  |
| Itally | 47 |  |
| United State | 97 |  |
| Australia | 82 |  |

**Figure 4:** Studies conducted across the globe on the prevalence of influenza viruses associated with wastewater.

**Notable Factors**

Natural and acquired host	Environmental persistence	Virus stability and transmissibility	Anthropogenic interventions

pH Temperature Salinity

Ottawa	Germany	America

Australia	America

**Figure 5**: A dendrogram that illustrates some of the notable factors that influence the transmission of influenza viruses in different regions.
