## Supplementary Table 1 for "A systematic review on the incidence of influenza viruses in wastewater matrices: Implications for Public Health"

**Table 1: Most productive authors in research related to the prevalence of Influenza viruses in wastewater milieu from 2012 to 2022 (Scopus and Google scholar)**

| **AUTHORS** | **TITLE** | **SOURCE TITLE** | **CITED BY** | **DOCUMENT TYPE** | **H-INDEX** | **COUNTRY** | **PUBLICATION DATE** |
| --- | --- | --- | --- | --- | --- | --- | --- |
| Bi *et al.,* | Simultaneous detection and mutation surveillance of SARS-CoV-2 and multiple respiratory viruses | Med | 9 | Review | 12 | United State | 2021 |
| Boehm *et al.* | Wastewater surveillance of human influenza, metapneumovirus, parainfluenza, respiratory syncytial virus (RSV), rhinovirus, and seasonal coronaviruses during the COVID-19 pandemic | Environmental Science and Health | 6 | Review | 12 | USA | 2022 |
| Brian *et al.* | A water-focused one-health approach for early detection and prevention of viral outbreaks | One Health | 43 | Article | 20 | United State | 2019 |
| Brisebois *et al.* | Human viral pathogens are pervasive in wastewater treatment center | Journal of Environmental Science | 62 | Article | 30 | China | 2018 |
| Dumke *et al.* | Simultaneous Detection of SARS-CoV-2 and Influenza Virus in Wastewater of Two Cities in Southeastern Germany, January to May 2022 | International Journal of Environmental Research and Public Health | 1 | Article | 24 | Germany | 2022 |
| Gaidet *et al.* | Understanding the ecological drivers of avian influenza virus infection in wildfowl: A continental-scale study across Africa | Processing of the Royal Society B: Biological Sciences | 109 | Article | 20 | Africa | 2012 |
| Ghernaout *et al.* | New insights towards disinfecting viruses | Journal of Water Reuse and Desalination | 15 | Article | 10 | Algeria | 2020 |
| Kevill *et al.* | Assessment of two types of passive sampler for the efficient recovery of SARS-CoV-2 and other viruses from wastewater | Science of the Total Environment | 3 | Article | 6 | United kingdom | 2022 |
| Mercier *et al.* | Wastewater surveillance of influenza activity: Early detection, surveillance, and subtyping in city and neighbourhood communities | Scientific Reports The preprint server for Health Science | 2 | Article | 6 | Canada | 2022 |
| O'Brien *et al.* | A water-focused one-health approach for early detection and prevention of viral outbreaks | One Health | 45 | Review | 20 | United State | 2019 |
| Ramos *et al.* | A robust, safe and scalable magnetic nanoparticles workflow for RNA extraction of pathogens from clinical and wastewater samples | Global Challenges | 10 | Article | 6 | Germany | 2021 |
| Ronnqvist *et al.* | Detection Method for Influenza Viruses in Water | International Journal of Environmental Research and Public Health | 11 | Article | 8 | Finland | 2012 |
| Rusinol *et al.* | Concentration methods for the quantification of coronavirus and other potentially pandemic enveloped virus from wastewater | Current Opinion in Environmental Science and Health | 65 | Review | 6 | Spain | 2020 |
| Silverman and Boehm | Systematic Review of the Persistence of Enveloped Viruses in Environmental Waters and Wastewater | Environmental Science & Technology Letters | 109 | Review | 12 | United State | 2021 |
| Sim *et al.* | Future perspectives of wastewater-based epidemiology: Monitoring infectious disease spread and resistance to the community level | Environmental International | 431 | Review | 25 | UK | 2020 |
| Teirlinck *et al.* | Annual report Surveillance of influenza and other respiratory infections in the Netherlands | Scientific Reports The preprint server for Health Science | 15 | Article | 18 | Netherlands | 2017 |
| Wigginton *et al.* | Emerging investigators series: the source and fate of pandemic viruses in the urban water cycle | Environmental Science :Water Research & Technology | 190 | Review | 20 | United State | 2015 |
| Wolfe *et al.* | Wastewater-Based Detection of Two Influenza Outbreaks | [Environmental Science and Technology Letters](https://www.scopus.com/sourceid/21100403504?origin=resultslist) | 6 | Article | 12 | United State | 2022 |
| Ye *et al.* | Survivability, Partitioning, and Recovery of Enveloped Viruses in Untreated Municipal Wastewater | Environmental Science and Technology | 356 | Article | 30 | United States | 2016 |
| Zhang *et al.* | Occurrence of various viruses and recent evidence of SARS-CoV-2 in wastewater systems | Journal of Hazardous Materials | 35 | Review | 24 | China | 2021 |
