## Supplementary Table 2 for "A systematic review on the incidence of influenza viruses in wastewater matrices: Implications for Public Health"

**Table 2**. Summary of the study details, sampling, and laboratory methodologies

| **Author (Year)** | **Country** | **Population Size** | **Number of Samples** | **Sampling**  **techniques** | **Sample type**  **(Locations)** | **PCR methods** |
| --- | --- | --- | --- | --- | --- | --- |
| Bi *et al.*(2021) | United State | Not mentioned | - | Grab and composite | Raw wastewater  (WWTPs) | RT-qPCR |
| Boehm *et al.*(2022) | USA | 1,500,000 | - | Grab | Raw wastewater  (WWTPs) | RT-qPCR |
| Brian *et al.*(2019) | United State | 290,000 | - | Composite | Wastewater  (WWTPs) | RT-qPCR |
| Brisebois *et al.* (2018) | China | Not mentioned | - | Composite | Raw and treated wastewater  (WWTPs) | q-PCR |
| Dumke *et al.* (2022) | Germany | - | 273 | Composite | Raw wastewater  (WWTPs) | RT-qPCR |
| Gaidet *et al.*(2012) | Africa | - | 8413 | Composite | WWTPs | RT-PCR |
| Ghernaout *et al.* (2020) | Algeria | - | - | Grab and composite | Raw and treated wastewater | RT-qPCR |
| Kevill *et al.* (2022) | United Kingdom | 40, 000 | 201 | Passive sampling | Raw wastewater | q-PCR |
| Mercier *et al.* (2022) | Canada | 910, 000 | - | 24-h Composite | Raw wastewater  (WWTPs) | RT-qPCR |
| O'Brien *et al.* (2019) | United State | 290, 000 | - | - | - | - |
| Ramos *et al.* (2021) | Germany | - | - | Composite | Clinical and raw wastewater samples | RT-qPCR |
| Ronnqvist *et al.*(2012) | Finland | - | - | Grab and composite | Wastewater, lake and river | Real time PCR |
| Rusinol *et al.* (2020) | Spain | - | - | composite | Raw wastewater | q-PCR |
| Silverman and Boehm (2021) | United State | 27 | 812 | Composite | Raw wastewater | RT-qPCR |
| Sim *et al.* (2020) | UK | - | - | Grab and composite | Treated and raw wastwater | RT-qPCR |
| Teirlinck *et al* (2017) | Netherland | 500, 000 | - | Grab and composite | Treated and raw wastwater | RT-qPCR |
| Wigginton *et al.*(2015) | United State | 398 | - | Grab and composite | Raw wastwater | RT- qPCR |
| Wolfe *et al.* (2022) | United State | 130, 000 | - | 24h composite | Raw wastewater | ddPCR |
| Ye *et al.*(2016) | United State | 115 000 | - | Grab | Untreated wastewater | RT- qPCR |
| Zhang *et al.* (2021) | China | - | - | Grab and composite | Treated and raw wastwater | RT-qPCR |
